# Clinical and immunological benefits of convalescent plasma therapy in severe COVID-19: insights from a single center open label randomised control trial

**DOI:** 10.1101/2020.11.25.20237883

**Authors:** Yogiraj Ray, Shekhar Ranjan Paul, Purbita Bandopadhyay, Ranit D’Rozario, Jafar Sarif, Abhishake Lahiri, Debaleena Bhowmik, Janani Srinivasa Vasudevan, Ranjeet Maurya, Akshay Kanakan, Sachin Sharma, Manish Kumar, Praveen Singh, Rammohan Roy, Kausik Chaudhury, Rajsekhar Maiti, Saugata Bagchi, Ayan Maiti, Md. Masoom Perwez, Abhinandan Mondal, Avinash Tewari, Samik Mandal, Arpan Roy, Moumita Saha, Durba Biswas, Chikam Maiti, Sayantan Chakraborty, Biswanath Sharma Sarkar, Anima Haldar, Bibhuti Saha, Shantanu Sengupta, Rajesh Pandey, Shilpak Chatterjee, Prasun Bhattacharya, Sandip Paul, Dipyaman Ganguly

## Abstract

**Introduction:** A single center open label phase II randomised control trial was done to assess the pathogen and host-intrinsic factors influencing clinical and immunological benefits of passive immunization using convalescent plasma therapy (CPT), in addition to standard of care (SOC) therapy in severe COVID-19 patients, as compared to patients only on SOC therapy.

**Methods:** Convalescent plasma was collected from patients recovered from COVID-19 following a screening protocol which also included measuring plasma anti SARS-CoV2 spike IgG content. Retrospectively, neutralizing antibody content was measured and proteome was characterized by LC-MS/MS for all convalescent plasma units that were transfused to patients. Severe COVID-19 patients with evidence for acute respiratory distress syndrome (ARDS) with PaO2/FiO2 ratio 100-300 (moderate ARDS) were recruited and randomised into two parallel arms of SOC and CPT, N=40 in each arm. Peripheral blood samples were collected on the day of enrolment (T1) followed by day3/4 (T2) and day 7 (T3). RT-PCR and sequencing was done for SARS-CoV2 RNA isolated from nasopharyngeal swabs collected at T1. A panel of cytokines and neutralizing antibody content were measured in plasma at all three timepoints. Patients were followed up for 30 days post-admission to assess the primary outcomes of all cause mortality and immunological correlates for clinical benefits.

**Results:** While across all age-groups no statistically significant clinical benefit was registered for patients in the CPT arm, significant immediate mitigation of hypoxia, reduction in hospital stay as well as survival benefit was recorded in severe COVID-19 patients with ARDS aged less than 67 years receiving convalescent plasma therapy. In addition to its neutralizing antibody content a prominent effect of convalescent plasma on attenuation of systemic cytokine levels possibly contributed to its benefits.

**Conclusion:** Precise targeting of severe COVID-19 patients is necessary for reaping the clinical benefits of convalescent plasma therapy.

**Clinical trial registration:** Clinical Trial Registry of India No. CTRI/2020/05/025209

## Introduction

The ongoing pandemic caused by the novel coronavirus SARS-CoV2 infection has already claimed more than 1.35 million lives, with close to 60 million documented infections worldwide. The acute respiratory disease caused by SARS-CoV2 infection, the coronavirus disease 2019 or COVID-19, present with a plethora of symptoms usually found to be spread over two distinct temporal phases in patients who are symptomatic. The symptoms in the initial milder phase variably include malaise, fatigue, fever, cough, loss of smell and taste and diarrhoea in some, mostly followed by recovery (Huang et al., 2020). But in a considerable fraction of patients this milder phase later on culminates in a more severe disease, characterized by gradually worsening hypoxemia requiring exogenous O2 supplementation. In some patients suffering from this severe disease a progression to acute respiratory distress syndrome (ARDS) is encountered, leading to untoward fatal outcomes in a number of them (Huang et al., 2020; WHO, 2020). Individuals having metabolic co-morbidities have been shown to have a predilection for COVID-19 disease severity (Lim et al., 2020). An aberrant hyper-activation of the immune system has been found to be associated with these severe symptoms, most notably characterized by by a systemic deluge of inflammatory cytokines or ‘cytokine storm’ (Laing et al., 2020; Arunachalam et al., 2020; Lucas et al., 2020).

Apart from the medical interventions aimed at mitigating symptomatologies, different therapeutic approaches are currently being explored, either by repurposing specific anti-viral agents, viz. remdesivir (Goldman et al., 2020), or by using corticosteroids to affect immunomodulation (Horby et al., 2020), to treat patients progressing to the severe disease. A number of patients also present with intravascular thrombosis and thus a role for prophylactic and therapeutic anticoagulation has also found place in the standard of care in severe patients (Billett et al., 2020). But in the absence of proven efficacy of any specific pathogen-targeted therapy, convalescent plasma transfusion is an age-old strategy for passive immunization, with the primary intention to supplement non-recovering patients with antibodies against specific pathogens (Rubin, 2020). Convalescent plasma therapy (CPT) has emerged as a widely tried strategy against COVID-19 too, having been explored in a large number of clinical trials all over the world (Agarwal et al., 2020; Liu et al., 2020; Joyner et al., 2020; Rubin, 2020; Li L et al., 2020). We report here the insights gathered from a single center open label phase II parallel arm randomised control trial done in a single center in Eastern India, done on patients suffering from severe COVID-19 disease with evidence for progressing to mild to moderate acute respiratory distress syndrome and identify the clinical and immunological benefits of convalescent plasma transfusion.

## Methods

### Ethical approval

The randomized control trial (RCT) on passive immunization with convalescent plasma therapy and all associated studies were done with informed consent from the patients according to the recommendations and ethical approval from the Institutional Review Boards of all the concerned institutions, viz. CSIR-Indian Institute of Chemical Biology, Kolkata, India (IICB/IRB/2020/3P), Medical College Hospital, Kolkata (MC/KOL/IEC/NON-SPON/710/04/2020), India and Infectious Disease & Beleghata General Hospital (ID & BG Hospital), Kolkata, India (IDBGH/Ethics/2429). The RCT was approved by Central Drugs Standard Control Organisation (CDSCO) under Directorate General of Health Services, Ministry of Health & Family Welfare, Govt. of India (approval no. CT/BP/09/2020) and registered with Clinical Trial Registry of India (CTRI, No. CTRI/2020/05/025209), under Indian Council of Medical Research, India.

### Collection of convalescent plasma

Convalescent donors were recruited and screened at the Department of Immunohematology and Blood Transfusion, Medical College Hospital, Kolkata, India. The inclusion criteria for donors were: age > 18 years, males or nulliparous female convalescent volunteers with history of being positive for SARS-CoV2 on RT-PCR, having weight > 55Kg, complete resolution of symptoms at least 28 days prior to donation, and a negative RT-PCR test for SARS-CoV2 before plasma donation. Consenting convalescent patients not fit to donate blood based on the history and examination, who have had transfusion of blood products in last one year were excluded from donation. A questionnaire was used to collect data on the disease course from all convalescent donors. On the screening day, peripheral blood samples were drawn for the following pre-donation tests: blood group (ABO grouping and Rh phenotyping) and antibody screening for clinically significant antibodies (Extended Rh, Kell, Duffy, Kidd, MNS, antibody screen positive donors were excluded), complete blood count including hemoglobin, hematocrit, platelet count, total and differential leucocyte count (Hb > 12.5g/dl, platelet count > 1,50,000 per microliter of blood and TLC within normal limits were included), screening for HIV, HBV and HCV, MP and syphilis by serology and ID-NAT for Hep B and C and HIV1 (all non-reactive donors by both tests were included), total serum protein (donors with total serum protein > 6gm/dl will be accepted, as per Drugs and Cosmetics (Second Amendment) Rules, 2020. For initial 18 convalescent donors pre-donation screening for anti-SARS-CoV2 spike protein IgG content of their plasma could not be done due to absence of dependable assay kits. Once it was available (Euroimmun) all donors were also pre-screened for anti-SARS-CoV2 spike IgG. For first 18 donors it was done retrospectively. All donated plasma were tested for their neutralizing antibody content using an in vitro surrogate neutralization kit (Tan et al., 2020). For prescreened donors a value of 1.5 for the ratio optical density between the sample and calibrator was taken as a cut-off for inclusion. A fraction of convalescent plasma sample was also characterized for their proteome using LC-MS/MS (described below). Plasmapheresis on eligible donors were done on a Haemonetics MCS+ Cell Separator. 400 ml of plasma was collected and aliquoted with sterile connections (Terumo TSCD) in two plasma bags containing 200ml each and cryostored at −80 deg C, until commissioned for transfusion in an ABO-matched recipient.

### Trial design

The inclusion criteria for recruitment of severe COVID-19 patients as recipients of convalescent plasma in this open label phase II randomized control trial were: consenting patients admitted with RT-PCR proven COVID-19 with severe disease (fever or suspected respiratory infection, plus one of the following; respiratory rate >30 breaths/min, severe respiratory distress, SpO2< 90% at room air) with mild ARDS, defined as patients having partial pressure of oxygen in the arterial blood (PaO2)/fraction of inspired oxygen (FiO2) ratio of 200-300 mmHg or moderate ARDS, defined as PaO2/FiO2 100-200 mmHg, not on mechanical ventilation. Pregnant or breastfeeding mothers, patients with age less than 18 years, patients participating in any other clinical trial, patients having any clinical condition precluding infusion of blood products were excluded. The trial was designed to recruit 40 patients in each arm. Accordingly, 80 patients were enrolled in to the trial at ID & BG Hospital, Kolkata, India, the first patient being recruited on May 31, 2020 and the last on October 12, 2020. Patients fulfilling the inclusion criteria were randomized into either the SOC arm receiving only the standard of care therapy, as per current advisory, or in to the CPT group to receive two consecutive doses of ABO-matched 200ml convalescent plasma on two consecutive days, the first transfusion being on the day of enrolment, in addition to standard of care. Nasopharyngeal swabs, fecal samples and peripheral blood in EDTA vials were collected on the day of enrollment (time point 1 or T1). Then on the 3^rd^ day or 4^th^ day post-enrollment (time point 2 or T2) and finally on the 7^th^ day post-enrollment (time point 3 or T3) peripheral blood samples were taken.

### Standard-of-care

Most patients infected with SARS-CoV2 at diagnosis received Hydroxychloroquine 400 mg BD on first day followed by 400 mg OD for four days, Azithromycin 500mg OD for 5 days, Ivermectin 12 mg OD for 5days and Doxycyclin 100 mg BD for 10 days. At the clinical trial site (ID & BG Hospital, Kolkata, India) standard-of-care (SOC) in all patients with evidence for ARDS were: O2 therapy as per requirement, either intravenous or oral corticosteroids, for patients with D-dimer <1000 Fibrinogen Equivalent Units (FEU) prophylactic anticoagulation and for patients with D-dimer >1000 ng/ml FEU therapeutic anticoagulation using either low molecular weight heparin or unfractionated heparin, appropriate broad-spectrum antibiotic therapy based on clinical, biochemical and microbiological assessment, appropriate anti-diabetic therapy to maintain blood sugar below 200mg/dl, anti-hypertensive agents, as per requirement, were used to maintain systolic blood pressure 100-140 mm of Hg, diastolic blood pressure at 70-90 mm of Hg and mean arterial pressure >65 mm of Hg. Awake proning for 6-8 hours/day was attempted in all patients with evidence for ARDS. O2 therapy was designed to maintain SpO2 >95% with successive deployment of higher efficiency devices as required, viz. nasal cannula, face mask, face mask with reservoir. Patients unable to maintain O2 saturation in blood (SpO2) above 90% with face mask with reservoir, high flow nasal cannula (HFNC) or in some cases mechanical ventilation (MV) were deployed. For the purpose of kinetic analysis of the SpO2/FiO2 ratio, a value of 89.99 was used for data-points where either HFNC or MV was in use. 1 patient in the SOC arm received Tocilizumab, none in the CPT arm. 13 patients in the SOC arm and 11 patients in the CPT arm received Remdesivir.

### Plasma cytokine analysis

Plasma isolated from peripheral blood of patients collected in EDTA vials. Plasma cytokine levels (pg/ml) were measured using the Bio-Plex Pro Human Cytokine Screening Panel 48-Plex Assay (Bio-Rad, Cat No. 12007283, for FGF basic, Eotaxin, G-CSF, GM-CSF, IFN-γ, IL-1β, IL-1RA, IL-1α, IL-2RA, IL-3, IL-12p40, IL-16, IL-2, IL-4, IL-5, IL-6, IL-7, IL-8, IL-9, GRO-α, HGF, IFN-α2, LIF, MCP-3, IL-10, IL-12p70, IL-13, IL-15, IL-17A, IP-10, MCP-1, MIG, β-NGF, SCF, SCGF-β, SDF-1α, MIP-1α, MIP-1β, PDGF-BB, RANTES, TNF-α, VEGF, CTACK, MIF, TRAIL, IL-18, M-CSF and TNF-β), using manufacturer’s protocol.

### RNA Isolation from COVID-19 Samples in TRIzol and RT-PCR

RNA from COVID-19 samples in TRIzol samples were extracted using chloroform-isopropanol method. qRT-PCR for SARS-CoV-2 detection was performed using the STANDARD M nCoV Real-Time Detection kit (Cat No. 11NCO10, SD Biosensor), approved by Indian Council of Medical Research (ICMR), as per manufacturer’s protocol. The RT-PCR was run on QuantStudio 6 Flex Real-Time PCR Systems (Applied Biosystems, Thermo Fisher Scientific). A cycle threshold (Ct-values) cut-offs mean value for both RdRp and E gene was considered as per SD biosensor’s manual for interpreting the results. CY5 labeled Internal Control is used as a positive control.

### SARS-CoV-2 Whole genome sequencing using Nanopore platform

In brief, 100ng total RNA was used for double-stranded cDNA synthesis by using Superscript IV (ThermoFisher Scientific, Cat.No. 18091050) for first strand cDNA synthesis followed by RNase H digestion of ssRNA and second strand synthesis by DNA polymerase-I large (Klenow) fragment (New England Biolabs, Cat. No. M0210S). Double stranded cDNA thus obtained was purified using AMPure XP beads (Beckman Coulter, Cat. No. A63881). SARS-CoV-2 genome was then amplified from 100ng of the purified cDNA following the ARTIC V3 primer protocol. Sequencing library preparation consisting of End Repair/dA tailing, Native Barcode Ligation, and Adapter Ligation was performed with 200ng of the multiplexed PCR amplicons according to Oxford Nanopore Technology (ONT) library preparation protocol-PCR tiling of COVID-19 virus (Version: PTC_9096_v109revE_06Feb2020). Sequencing in sets of 24 barcoded samples was performed on MinION Mk1B platform by ONT.

### Nanopore analysis

The ARTIC end to end pipeline was used for the analysis of MinION raw fast5 files up to the variant calling. Raw fast5 files of samples were basecalled and demultiplexed using Guppy basecaller that uses the basecalling algorithms of Oxford Nanopore Technologies (https://community.nanoporetech.com) with phred quality cut-off score >7 on GPU-linux accelerated computing machine. Reads having phred quality score less than 7 were discarded to filter the low-quality reads. The resulting demultilexed fastq were normalized by read length of 300-500 (approximate size of amplicons) for further downstream analysis and aligned to the SARS-CoV-2 reference (MN908947.3) using the aligner Minimap2 v2.17 (Li H, 2018). Nanopolish were used to index raw fast5 files for variant calling from the minimap output files (Loman et al., 2015). To create consensus fasta, bcftools v1.8 was used over normalized minimap2 output.

### Phylogenetic reconstruction

The assembled SARS-CoV-2 genomes were aligned using MUSCLE aligner in default mode using the software UGENE v34 (Edgar, 2004; Okonechnikov et al., 2012). The phylogenetic tree construction was performed using the Maximum Likelihood method. Visualization and further editing of the tree was done in FigTree 1.4.4 (Rambaut, 2012).

### SARS-CoV-2 Surrogate Virus Neutralization Assay

Neutralizing antibodies against SARS-CoV-2 in human plasma samples from peripheral blood of convalescent donors were detected using GeneScript SARS-CoV-2 Surrogate Virus Neutralization kit (Cat no-L00847). Assay was performed according to manufacturer’s protocol. Plasma samples, and provided positive and negative controls were diluted at a ratio of 1:10 with the sample dilution buffer. Presence of SARS-CoV-2 neutralizing antibodies in the plasma samples results in inhibition of interaction between HRP-RBD and plate-bound human ACE2 protein, and subsequent development of color, assay results are interpreted as inhibition rate of assay reaction. The neutralizing antibody content was measured for all convalescent plasma samples as well as for recipients at T1, T2 and T3.

### ELISA for anti SARS-CoV-2 IgG

Levels of Immunoglobulin G (IgG) specific for SARS-CoV-2 in the plasma isolated from peripheral blood of convalescent donors were detected using EUROIMMUN Anti-SARS-CoV-2 (IgG) Elisa kit (Cat No-EI 2606-9601 G). This assay provides semiquantitative estimation of IgG levels against SARS-CoV-2 spike protein. Assay was performed according to manufacturer’s protocol. Presence of anti SARS-CoV-2 IgG antibodies in the plasma was measured using the following formula: Ratio = Extinction of the control or patient samples/Extinction of calibrator (Ratio ≥ 1.1 is interpreted as positive).

### Proteomics analysis of convalescent plasma

From each sample, 10 µl of plasma was taken in a fresh 1.5 ml micro-centrifuge tube and diluted to 100 µl with phosphate buffer (1X PBS). Rapid protein precipitation was performed for these samples by addition of 400 µl of acetone and incubated at room temperature for 2 minutes followed by centrifugation at 10000 g for 5 minutes (as described by https://doi.org/10.1021/acs.jproteome.9b00867). After removal of supernatant, pellets were air dried and resuspended in 100 mM Tris-HCl buffer (pH 8.5).Protein estimation was performed for each samples using the Bradford assay (Sigma-Aldrich, USA). For proteomics analysis, 20 µg of protein from each sample was reduced by addition of 25 mM of dithiothreitol (Sigma-Aldrich, USA) and incubated at 56 ºC for 25 minutes. Cysteine alkylation was performed by addition of 55 mM iodoacetamide (Sigma-Aldrich, USA) and incubated in dark for 20 minutes. Samples were subjected to trypsin (sequencing grade, Promega) digestion at an enzyme to substrate ratio of 1:10 for 18 hours at 37 ºC. Reaction was terminated by addition of 0.1% formic acid and dried under vacuum. Peptide clean up was performed using Oasis HLB 1cc Vac cartridge (Waters). DIA-SWATH analysis for samples were performed on a quadrupole-TOF hybrid mass spectrometer (TripleTOF 6600, Sciex, USA) coupled to a nano-LC system (Eksigent NanoLC-425). For each sample, 4 μg of these peptides were loaded on a trap-column(ChromXP C18CL 5 μm 120 Å, Eksigent) where desalting was performed using 0.1% formic acid in water with a flow rate of 10 μl per minutes for10 min. Peptides were then separated on a reverse-phase C18analytical column (ChromXP C18, 3 μm 120 Å, Eksigent) in a 57 minutes gradient of buffer A (0.1%formic acid in water) and buffer B (0.1% formic in acetonitrile) at a flow rate of 5 μl/minute. Buffer B was slowly increased from 3% at 0 minute to 25% in 38 minutes, further increased to 32% in next 5 minutes and ramped to 80% buffer B in next 2 minutes. In 0.5 min, buffer B was increased to 90% and column was washed for 2.5 minutes, buffer B was brought to initial 3% in next 1 minute and column was reconditioned for next 8 minutes. A method with 100 precursor isolation windows were defined based on precursor m/z frequencies using the SWATH Variable Window Calculator (Sciex), with a minimum window of 5 m/z. The accumulation time was set to 250 msec for the MS scan (400–1250 m/z) and 25 msec for the MS/MS scans (100–1500 m/z). Rolling collision energies were applied for each window based on the m/z range of each SWATH and a charge 2+ ion, with a collision energy spread of five. The total cycle time was 2.8 sec. SWATH data was processed using the SWATH Acquisition MicroApp 2.0.1 in PeakView 2.2 Software. An in-house protein spectral-ion library file (.group) was imported with specified maximum 251 proteins and shared peptides were excluded. SWATH run files were added and retention time alignment was performed using peptides from abundant proteins. The processing settings for peak extraction were: at least 10 peptides per protein, 5 transitions per peptide, >95% peptide confidence threshold, 1 %FDR. XIC extraction window was set to 55 min with 75 ppm XIC Width. All information was exported in the form of MarkerView (.mrkw) files. In MarkerView1.2.1, data normalisation was performed using total area sum normalisation and exported to excel.

### Co-occurrence analyses

Co-occurrence among each pair of cytokines was calculated using Spearman correlation (r) and corresponding p-value of the correlation was measured using a t-distribution. Absolute values of the cytokines were used for the calculation of correlation network and threshold was set to r>=0.7, p<0.01 for the complete set of cytokines from SOC (n=40) and CPT (n=39) groups. All calculations were done using the ‘Hmisc’ R package and finally converted to a network file using the ‘igraph’ R package. Visualisation of the network was performed using Cytoscape 3.7.2. Each cytokine was color coded and node size was set proportional to the fold change of median as compared to the same cytokine in the mild datasets.

### Statistical analyses

All statistical analyses, as depicted in the results as well in appropriate figure legends, were performed using R and in some cases using Graphpad Prism 8 or STatistica64 (StatSoft).

## Results

### Recruitment of convalescent donors and characterization of antibody response

61 convalescent individuals (female:N:12, Age: 26 ± 2.98 years; male: N:49, Age: 35.37 ± 9.06 years) who recovered from COVID-19 at least 28 days prior to screening, were screened for eligibility for plasma donation. 46 donors were found eligible for plasmapheresis. All of them were screened at 40-80 days after they were first tested positive on RT-PCR for SARS-CoV2 (Figure 1A). The nature of their disease course were assessed to be between 1-5 on the WHO Clinical Progression score for with the majority having suffered from a mild symptomatic disease.

**Figure 1.**
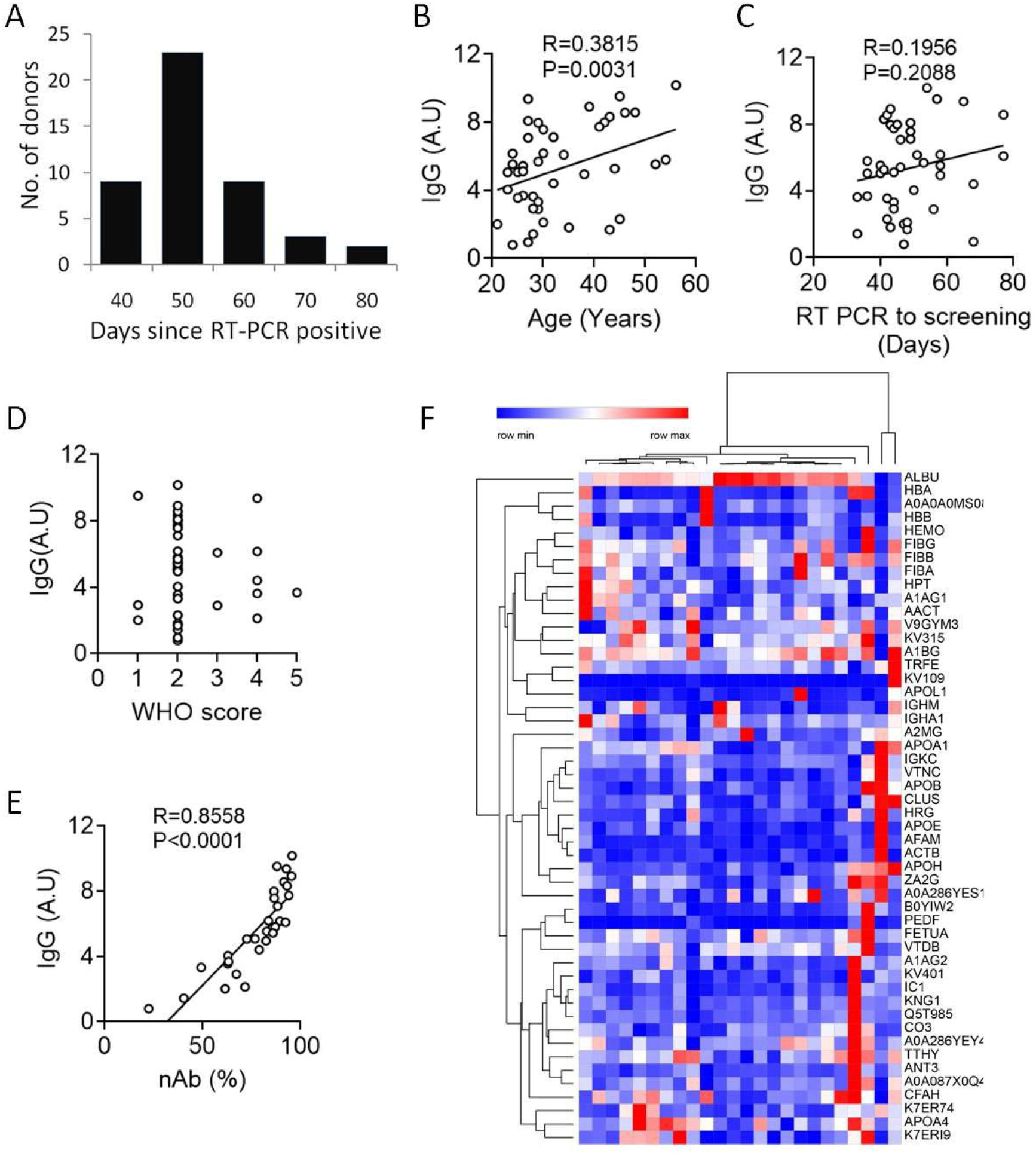
Convalescent donor characteristics. **(A)** Day since being tested positive for SARS-CoV2 on RT-PCR are shown for all convalescent donors. **(B)** Correlation between the anti SARS-CoV2 sike protein IgG content of plasma and age of the convalescent donors is shown. **(C)** Correlation between the anti SARS-CoV2 spike protein IgG content of plasma and day since the patients were tested positive on RT-PCR is shown. **(D)** Distribution of the convalescent donor cohort for the WHO Clinical Progression Score for their disease courses are plotted against the anti SARS-CoV2 spike protein IgG content of plasma. **(E)** Correlation between the anti SARS-CoV2 spike protein IgG content of plasma and the efficiency of plasma to neutralize the SARS-CoV2 spike protein RBD-ACE2 interaction is shown. **(F)** A heat map showing correlative clusters of 50 proteins present in convalescent donor plasma that show highest variance among donors. Pearson correlation was computed and the R and P values are noted when significant correlation is found.

On measuring the anti-SARS-CoV2 spike protein IgG content of plasma a significant correlation was found with age of the donors, with increasing age the a more robust humoral response and higher IgG content was noted (Figure 1B). Interestingly in this small cohort of convalescent donors no significant trend in terms of correlation between the time passed since positive RT-PCR test and SARS-COV-specific IgG content of plasma (Figure 1C). The IgG content was also not correlated with the WHO clinical progression score for the COVID-19 disease course that the donors reported (Figure 1D), although most of the donors had very similar milder disease course in our study. Notably a very strong correlation was noted between the anti-SARS-CoV2 spike IgG content of plasma and the content of neutralizing antibody (nAb) measured in an in vitro spike RBD-ACE2 interaction-based surrogate neutralization assay (Figure 1E). A proteomic analysis of the convalescent plasma was also performed using mass spectrometry for characterization of the major protein components of plasma. Proteins registering most variance among donors, in terms of the semiquantitative measure of area under curve, are listed (Figure 1F).

### Inter-arm patient characteristics with viral load and humoral response

80 patients (Female: N: 23, age: 61.43±11.33 years; Male: N: 57, age: 61.36±12.17 years), fulfilling the inclusion criteria were recruited into the trial and randomized into either standard of care (SOC) arm or the convalescent plasma therapy (CPT) arm. Age of the patients between two parallel arms was not significantly different (Figure 2A). The number of days since hospital admission at enrolment was 3.85±2.63 days for SOC arm and 4.2±2.21 days for CPT arm, showing no significant difference. Most of the patients recruited were suffering from moderate ARDS at the time of recruitment with a mean SpO2/FiO2 ratio of 108.38 on the day of enrolment for SOC arm and 111.43 for the CPT arm. 13 female patients and 27 male patients were randomized into the SOC arm. 10 female patients and 30 male patients were randomized into the CPT arm. All patients recruited in CPT arm received two transfusions of 200ml ABO-matched convalescent plasma on two successive days, first one being on the day on enrolment, except for one patient who succumbed before he could be transfused with second unit, but not due to any adverse effect that could be temporally linked to the first plasma transfusion. Transfusion-related adverse effects were reported in none of the patients in CPT arm.

**Figure 2.**
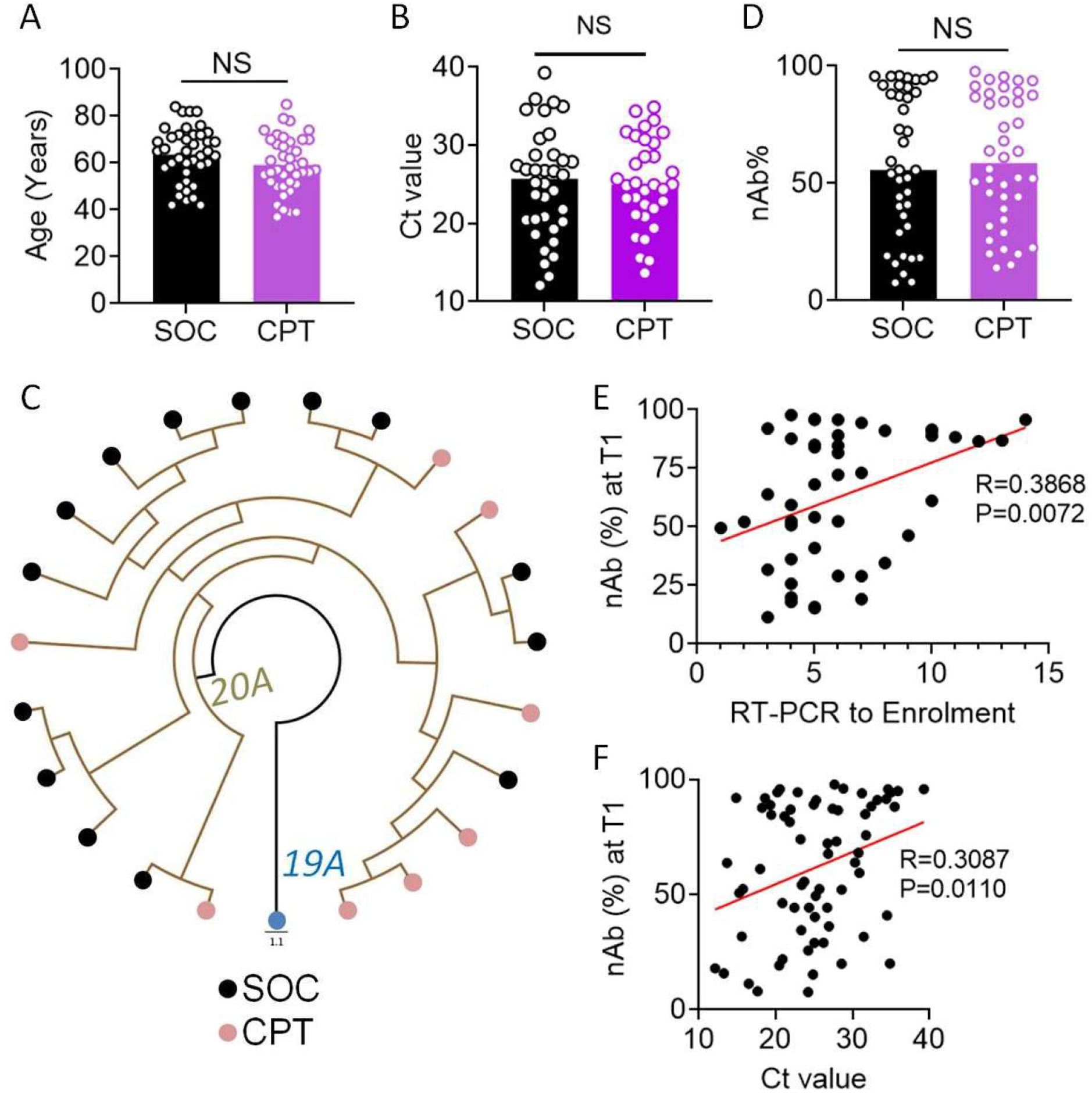
Key host and pathogen characteristics. **(A)** Age distribution of patients are compared between SOC and CPT arms. **(B)** CT values obtained from SARS-CoV2 RT-PCR from nasopharyngeal swabs collected on the day of enrolment (T1) are compared between SOC and CPT arms. **(C)** The whole genome phylogenetic tree of SARS-CoV2 viral isolates from SOC and CPT groups from clade 20A. Black and purple tips represent the samples from SOC and CPT groups respectively whereas blue tip represents the reference strain from Wuhan, China. **(D)** Neutralizing activity of plasma from patients collected at T1 is compared between SOC and CPT arms. **(E)** Correlation between the neutralizing antibody content of plasma at T1 and day since the patients were tested positive on RT-PCR is shown. **(F)** Correlation between the neutralizing antibody content of plasma at T1 and CT values obtained from SARS-CoV2 RT-PCR from nasopharyngeal swabs collected at T1 is shown. For all comparisons unpaired T tests were performed. Pearson correlation was computed and the R and P values are noted when significant correlation is found.

Viral loads at the day of enrolment (T1) were comparable between patients in the two arms (Figure 2B). Viral isolates were sequenced from nasopharyngeal swabs collected from 14 patients in the SOC arm and 8 patients in the CPT arm. There was no notable difference in viral clade representation among these patients recruited into two arms. All 22 patients showed infection with SARS-CoV2 clade 20A (Figure 2C). The neutralizing antibody content of plasma at T1 was also not significantly different between SOC and CPT (Figure 2D). A significant correlation between the neutralizing antibody content of plasma was noted with number of days passed since the patients first got tested positive for SARS-CoV2 on RT-PCR (Figure 2E). Across all patients, from both arms, a significant negative correlation between the neutralizing antibody content of plasma at T1 and concomitant viral load was found, as expected (Figure 2F).

### Greater attenuation of the cytokine storm with convalescent plasma therapy

The severe COVID-19 patients have been found by previous studies to experience a systemic immune hyper-activation characterized by a cytokine storm. We had previously reported nature and dimension of the cytokine storm in a fraction of these patients, comparing them to patients suffering from mild COVID-19 disease (Bandopadhyay et al, 2020). On measuring plasma abundance of a panel of 48 cytokines in patients from both arms we found that correlative nature and magnitude of the individual components of the cytokine storm were not notably different at T1 in a correlative network analysis (Figure 3A). Data from a panel of 36 cytokines were included in all analyses selected based on their measurable plasma abundance in at least 70% of the patients. The magnitude of plasma abundance was computed in comparison with median abundance of individual cytokines in patients having mild COVID-19 disease reported earlier (Bandopadhyay, 2020). As reported in this earlier study done in a smaller fraction of these patients, a more significant attenuation of the systemic deluge of cytokines at T2 were noted in patients in the CPT arm, both in terms of calming down of the correlative upregulation and reduction in the median abundance of major pathogenically significant cytokines (Figure 3B), as well as in terms of number of patients in CPT arm registering such a change (Figure 4).

**Figure 3.**
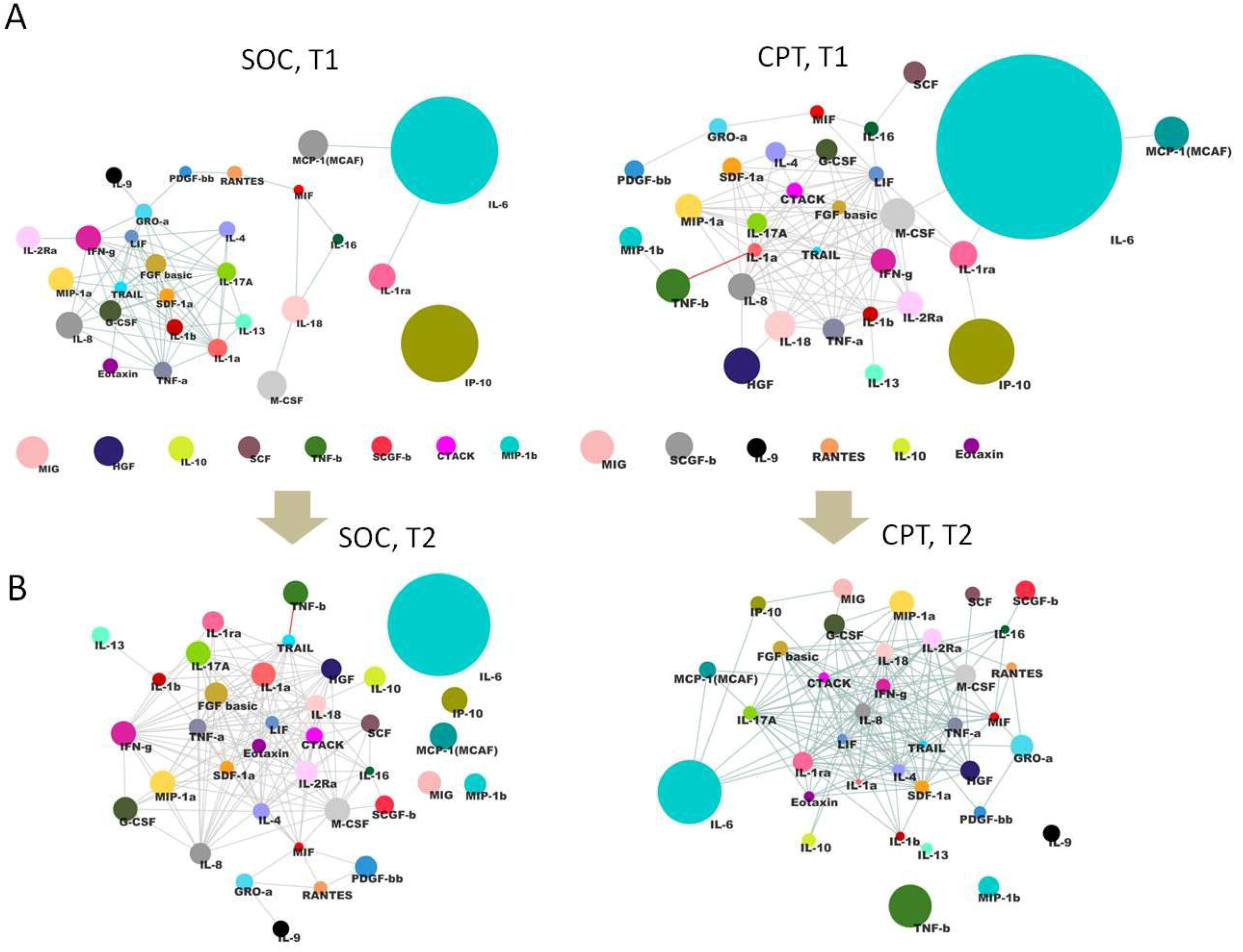
Cytokine expression correlation matrix. **(A)** and **(B)** describe the correlation network for 36 cytokines measured in plasma of severe COVID-19 patients, from both SOC and CPT group at T1 and T2 time points respectively. The diameter of the nodes represents extent of enriched abundance compared against a median value derived from patients with mild disease. Edges are shown only for Spearman correlation ρ value of 0.7 and above.

**Figure 4.**
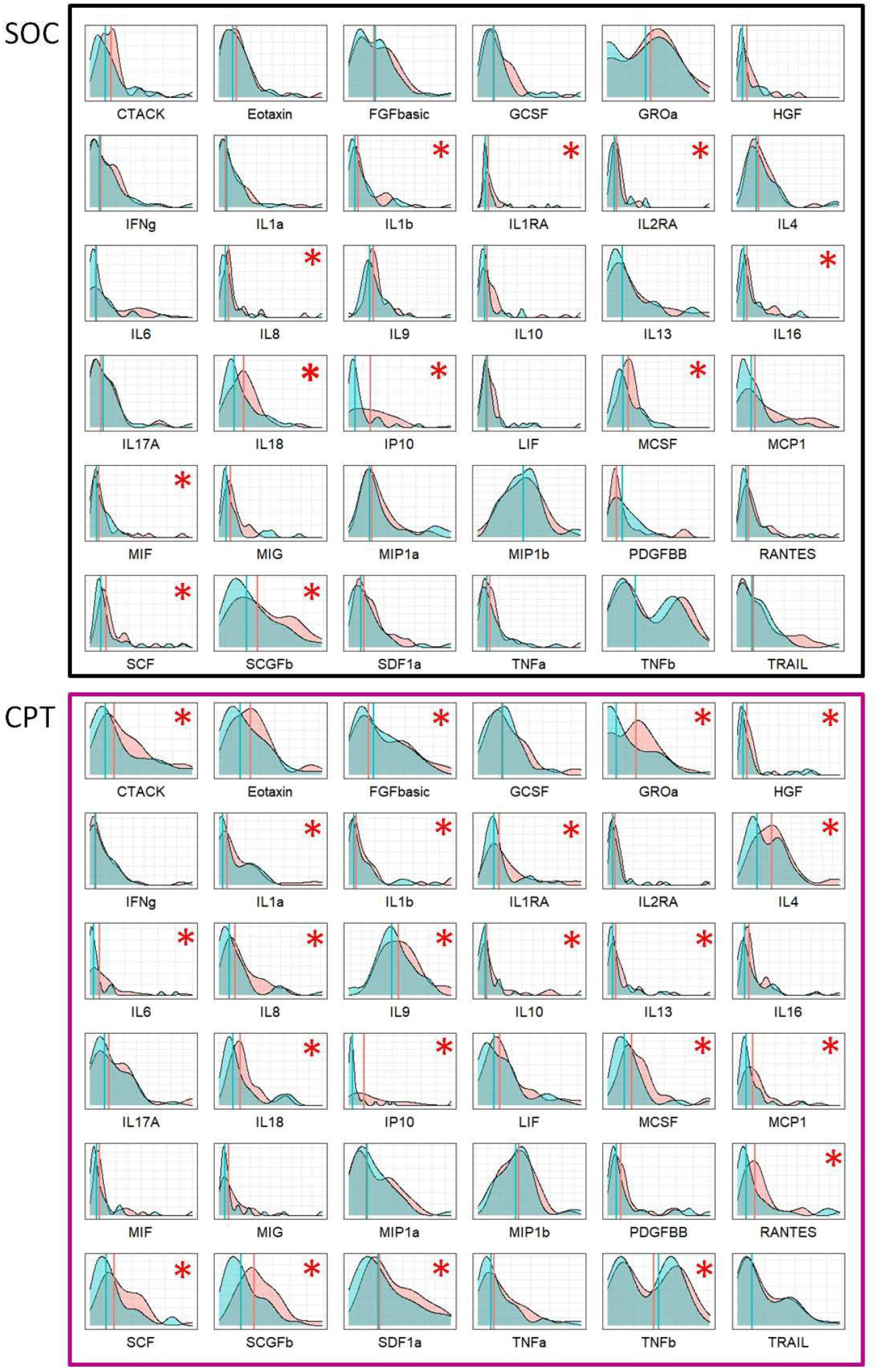
Distribution of cytokine expression levels in the patient cohort. Density histogram of plasma abundance levels of 36 cytokines at T1 and T2 time points. Upper panel represents the change in population distribution of cytokine abundance for SOC arm from T1 to T2 time point and lower panel represents the same for CPT arm. The pink and blue colour indicates the distribution profiles of indicated cytokines at T1 and T2 time points respectively. The red stars are marking significant differences in median values between T1 and T2.

### Assessment of clinical benefits of convalescent plasma therapy

On analysing the primary outcome of all cause mortality at 30 days, across all patients in two arms, we found no significant benefit in the intervention arm, either in terms of mitigation of hypoxia as represented by kinetics of SpO2/FiO2 ratio (S/F ratio) over 10 days following enrolment (Figure 5A), survival (Figure 5B, Mantel-Haenszel Hazard Ratio 0.6731, 95% confidence interval 0.3010-1.505, with a P value of 0.3424 on Mantel-Cox Log-rank test), duration of hospital stay since the day of enrolment (Figure 5C, median of 17 days for SOC vs 13 days for CPT arm, P value of 0.098 on Mantel-Cox Log-rank test) or duration of hospital stay since admission (Figure 5D, median of 23 days for SOC vs 17 days for CPT arm, P value of 0.0797 on Mantel-Cox Log-rank test).

**Figure 5.**
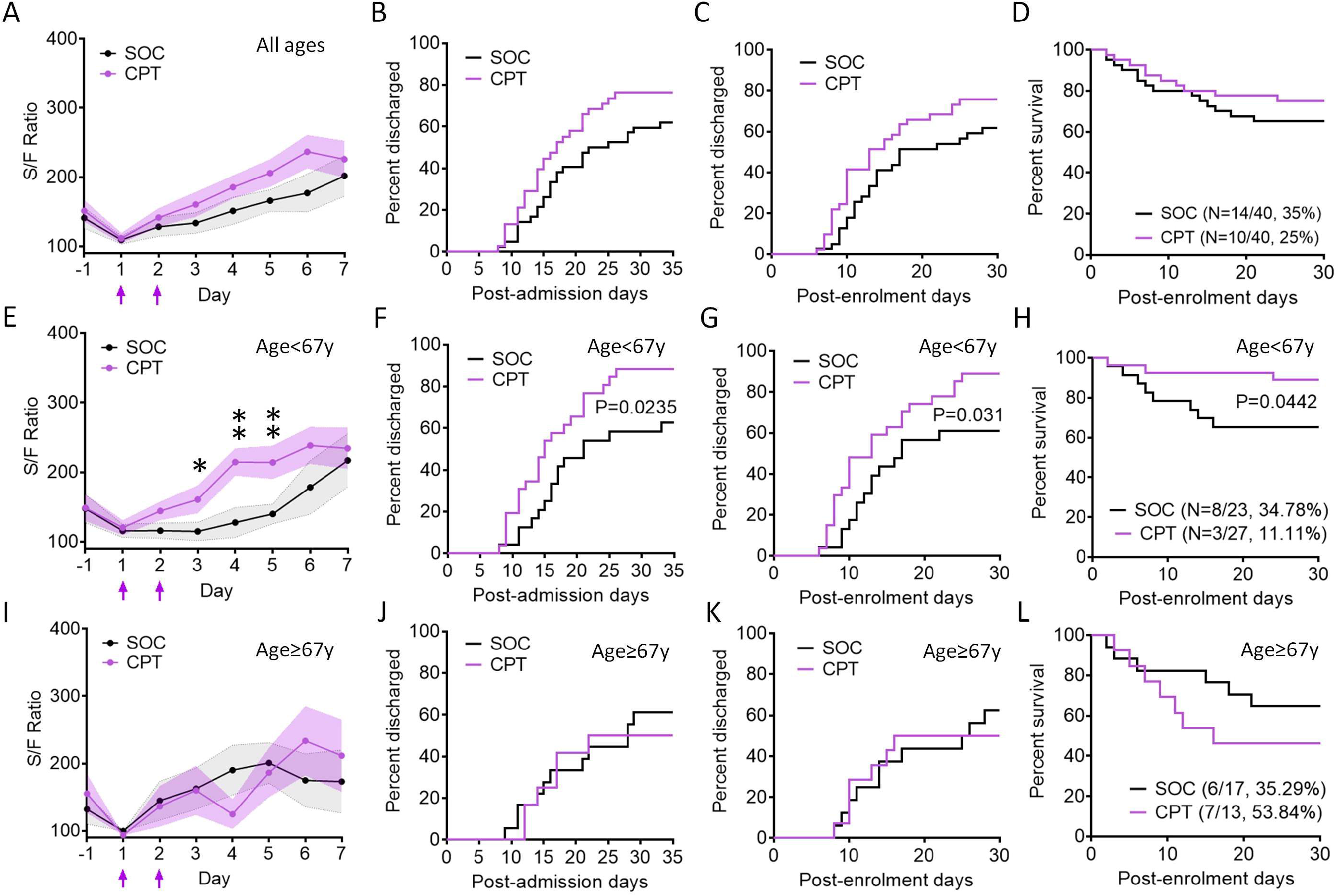
Clinical outcomes among patients in the two arms of the randomised control trial. **(A), (E) & (I)** The ratio between saturation of O2 in blood (SpO2) and fraction of O2 received (FiO2), or S/F ratio is plotted for patients in SOC (black line) and CPT (purple line) arms from the day before enrolment till th 7^th^ day post-enrolment, among all age-groups (A), for patients aged <67 years (E) and for patients aged ≥67 years (I). Purple arrows indicate the days when convalescent plasma was transfused. 95% confidence interval is shown for each group. * P<0.05, **P<0.005, from unpaired T tests. **(B), (F) & (J)** Total hospital stay duration of the patients from both arms are plotted in an ascending Kaplan-Meier curve, for all age groups (B), for patients aged <67 years (F) and for patients aged ≥67 years (J). Deaths and non-remission at day 35 post-admission were censored. **(C), (G) & (K)** Hospital stay duration of the patients from both arms since the day of enrolment are plotted in an ascending Kaplan-Meier curve, for all age groups (C), for patients aged <67 years (G) and for patients aged ≥67 years (K). Deaths and non-remission at day 30 post-enrolment were censored. **(D), (H) & (L)** Survival of patients in the two arms from the day of enrolment till day 30 post-enrolment are compared in a Kaplan-Meier curve, for all age groups (D), for patients aged <67 years (H) and for patients aged ≥67 years (L). Surviving patients were censored on day 30 post-enrolment. For all outcomes Mantel Cox log rank test was performed and corresponding P values are only shown for statistically significant differences.

Then we performed a sub-class analysis on the clinical outcome data taking different age-groups into consideration, with an aim to find an age-threshold which registers minimum hazard ratio but retains statistical power in this study with a relatively small size of the cohort. On this analysis, with an age threshold of <67 years significant differences in terms of clinical outcomes between two arms were noted. For patients with aged <67 years, a significant early mitigation of hypoxia was noted in response to convalescent plasma therapy (Figure 5E). Nevertheless, for patients aged <67 years a significant survival benefit was also noted in the CPT arm (Figure 5F, Mantel-Haenszel Hazard Ratio 0.2915, 95% confidence interval 0.08773-0.9685, with a P value of 0.0442 on Mantel-Cox Log-rank test). Also the surviving patients registered early remissions in terms of duration of hospital stay since the day of enrolment (Figure 5G, median of 17 days for SOC vs 13 days for CPT arm, P value of 0.031 on Mantel-Cox Log-rank test) or duration of hospital stay since admission (Figure 5H, median of 21 days for SOC vs 15 days for CPT arm, P value of 0.0235 on Mantel-Cox Log-rank test). For patients aged ≥67 years there was no significant difference in terms of either mitigation of hypoxia, survival or time to remission between the two arms (Figure 5I-L).

### Discussion

The open label randomised control trial for passive immunization of severe COVID-19 patients with convalescent plasma therapy adds to the growing literature on these trials of different designs and sample sizes. The present RCT was done in a low clinical resource setting in a single center. The clinical outcome comparisons revealed a significant benefit registered in severe COVID-19 patients, most of whom had progressed to moderate acute respiratory syndrome, and were aged less than 67 years. The trial also found a significantly early mitigation of hypoxia among patients in the same age-group receiving CPT and reaping the relative survival benefit. Of note here the patients in the SOC group who belonged to the same age-group also could catch up on this by day 6 post-enrolment. This may represent abrogation of the relative benefit offered by convalescent plasma with time since transfusion. Whether additional transfusions can further maintain this relative benefit resulting in faster remissions and higher survival remains to be explored. Another important revelation of this trial had been the prominent anti-inflammatory effects of CPT, which may also underlie the clinical benefits registered. Further explorations are warranted to validate the anti-inflammatory effect in larger cohorts and to identify non-immunoglobulin components of convalescent plasma responsible for these anti-inflammatory effects.

The major limitation of the present trial had been a small sample size, which perhaps prevented the trial from discerning the relative clinical benefits across all age-groups. Nevertheless the significant benefit registered in the <67 year age-group points to a detrimental effect of an aging immune system on disease remission as well as eventual outcomes in severe COVID-19. The biology underlying this will be of great interest in subsequent mechanistic studies as well as in future trials with specific monoclonal antibodies being developed for therapeutic usage in COVID-19.

## Data Availability

All data used in the manuscript are available with the corresponding author.

## Acknowledgements

D.G. acknowledges funding for the RCT and associated immune monitoring studies from Council of Scientitific Industrial Research (CSIR), Govt. of India (MLP-129); R.P. acknowledge funding from CSIR (MLP-2005) and Fondation Botnar. D.G. holds a Swarnajayanti Fellowship from Department of Science & Technology (DST), Govt. of India, S.P. holds a Ramanujan Fellowship from Science & Engineering Research Board (SERB), Govt. of India, S.C. holds an Intermediate Fellowship from DBT-Welcome Trust India Alliance and R.P. holds a Ramalingaswami Fellowship from Department of Biotechnology (DBT), Govt. of India. P.B. and A.L. are supported by Senior Research Fellowship from CSIR, R.D. and J.S. are supported by Junior Research Fellowships from University Grants Commission, India and D.B. is supported by Senior Research Fellowship from Indian Council of Medical Research, India. Authors express their gratitude to Abhijit Chowdhury and Anurag Agrawal for their valuable guidance while conceiving the trial and Mohd. Faruq for help with RT-PCR experiments.

## Author contributions

D.G. conceptualized the study. D.G. and Y.R. designed the study protocol. P.B. and R.D. did the plasma cytokine analysis. JS performed serological studies. A.L. contributed to the analysis and computation. Y.R. and S.R.P. recruited patients, maintained clinical data and supervised clinical management. R.R., R.M., K.C., S.B., Ay.M., M.M.R., A.T., Av.M. S.M., B.S.S., A.H. and B.S. contributed to patient management. J.S.V., R.M., A.K. and R.P. did the RT-PCR for SARS-CoV2 and sequencing viral RNA. D.B. contributed in analysis of viral genome and clade identification. P.S. and S.S. did the proteomics experiments. S.C. contributed in immunological studies. S.P. and P.B. recruited convalescent donors, D.B., C.M. and P.B. designed the donor selection guidelines, performed donor screening, apheresis and biobanking of convalescent plasma. D.G. wrote the manuscript with inputs from other authors. All authors approved the manuscript.

